# Directed effective connectivity between the SCN-containing hypothalamus and pineal gland varies by chronotype in bipolar disorder

**DOI:** 10.64898/2026.09.24.26363889

**Authors:** Ida Mehrdadi, Sonja Kuster, Jonas Rohrer, Erich Seifritz, Céline Chatelain, Gabrielle Chan, Philipp Homan, Marlene Tahedl

## Abstract

Circadian disruption is a transdiagnostic feature of psychiatric illness, particularly in bipolar disorder (BD), where evening chronotype is linked to greater symptom severity. The suprachiasmatic nucleus (SCN) times pineal gland (PG) melatonin synthesis through a well-described feedforward pathway. Moreover, melatonin can also phase-shift the SCN-pacemaker, consistent with a functional feedback limb. Directed effective connectivity between the SCN-containing hypothalamus and the PG has not been estimated in human neuroimaging, and it is unknown whether that directed coupling varies by psychiatric diagnosis or chronotype. We applied cross-spectral dynamic causal modeling (DCM) to resting-state fMRI data in 431 UK Biobank (healthy controls [HC, N=136], BD [N=78], major depressive disorder [MDD, N=154] and psychotic disorders [PY, N=63]). Random-effects Bayesian model selection compared a fully connected model with reduced archiature and, separately, feedforward-only (SCN→PG) vs feedback-only (PG→SCN) models. A per-subject log Bayes factor difference (dF) indexed directionality preference and was tested against diagnosis, chronotype and their interaction using ANCOVA. The fully connected model was strongly preferred (posterior probability=97.8%, protected exceedance probability [PXP]=1.00). Feedforward architecture dominated overall (70% vs 30%, PXP=1.00). Diagnosis did not predict dF (*p*=.403), but chronotype did (*p*=.036), and there was a significant diagnosis-by-chronotype interaction (*p*<.001). Feedforward dominance was strongest in evening chronotypes (p<.001). Model evidence favored a fully connected architecture with overall feedforward dominance. That preference varied by chronotype and, qualified by the interaction, was strongest in BD evening chronotypes. The pattern supports BD-evening chronotype as a high-risk subgroup and argues for routine chronotype assessment in BD care.

## Introduction

Circadian rhythm disruption is a prominent transdiagnostic feature across psychiatric disorders [1–3]. It is particularly well-documented in bipolar disorder (BD) [4], where it manifests as abnormal sleep-wake cycles, evening chronotype preference, and misalignment between endogenous timing and external zeitgebers [5–7]. Evening chronotype in BD has been associated with more severe symptoms, higher rates of mood episodes, increased comorbidity, and poorer long-term outcomes, suggesting it represents a distinct high-risk subtype [8, 9]. Changes in the structure and function of the suprachiasmatic nucleus (SCN), the main circadian pacemaker, and its surrounding hypothalamic region, as well as disruptions in melatonin secretion – the primary hormonal output of the circadian system, synthesized in the pineal gland (PG) – are linked to BD pathophysiology [10–13]. In our prior multimodal work, some to these signatures showed lithium-associated attenuation [14].

The SCN orchestrates the circadian rhythm of melatonin production in the PG through a multisynaptic sympathetic pathway [15–18]. Classically, this control was considered predominantly feedforward: the SCN provides tonic inhibition of pineal melatonin synthesis during the biological day via GABAergic suppression of sympathetic outflow and releases this inhibition at night, permitting norepinephrine-driven melatonin synthesis [19–23]. High-affinity melatonin receptors (MT1 and MT2) on SCN neurons provide a substrate for feedback [24, 25], and exogenous melatonin phase-shifts the human pacemaker, according to a well-described phase-response curve [26–28]. Melatonin acutely inhibits SCN neuronal firing and can phase-shift the pacemaker, consistent with a functional feedback limb that modulates circadian timing and entrainment [19, 29–33].

What has not been estimated in humans is directed effective connectivity between the SCN-containing hypothalamus and the PG using neuroimaging. Dynamic Causal Modeling (DCM) of resting-state fMRI data provides a Bayesian framework for comparing competing models of such directed, “effective” connectivity and quantifying their relative evidence [34, 35]. In the present study, we tested three a priori hypotheses in a large transdiagnostic cohort comprising healthy controls (HC), patients with BD, major depressive disorder (MDD), and psychotic disorders (PY). First, we hypothesized that a fully connected bidirectional model (SCN-containing hypothalamic subunit ⇄ PG) would be preferred over simpler architectures. Second, we expected overall dominance of feedforward (from the SCN-containing hypothalamic region to the pineal gland) models, consistent with the SCN region’s role as the primary driver of the system [15]. Third, we hypothesized that directionality preference would be modulated by diagnosis and/or chronotype, with the strongest effects in BD evening chronotypes – potentially reflecting greater dominance or reduced flexibility of SCN-gating in this high-risk subgroup, consistent with our previous findings [14].

To test these hypotheses, we applied DCM to resting-state fMRI data from 431 participants (comprising BD, MDD, PY, and HC) while controlling for age, sex, handedness, medication regimen, and time of day. Directionality preference was quantified as the difference in model evidence between feedback-only and feedforward-only models and was examined as a function of diagnostic group and morningness-eveningness chronotype. Analyses respected the anatomical distinction between left and right SCN-containing subregions [36].

## Methods and Materials

### Dataset

We utilized samples from the UK Biobank [37]. We included *n* = 603 participants for preprocessing. Selection criteria from Tahedl, Rohrer, Seifritz, et al. (2026) were used for preprocessing inclusion. In brief, participants were selected from UK Biobank participants with a baseline T1w MRI scan, where ICD-10 codes were used to identify patient subgroups: BD (F31x, *n* = 113), MDD (F32x, *n* = 205), PY (F2x, *n* = 91). Initially, 2595 MDD participants were available. The sample was reduced to better match other groups by excluding psychiatric/neurological comorbidities and requiring antidepressant use. HC were defined as those with no registered ICD-10 diagnosis and were randomly subsampled to match mean age and sex of the BD group, the main group of interest to this study. Of the 603 participants screened, 431 were included for final analysis (see Supplementary Figure 1 for a flowchart of participant inclusion/exclusion criteria). Groups were further subdivided into evening and morning chronotype using the UK Biobank field 1180 - morning or evening person (chronotype), a validated self-report measure [38]. The questionnaire asks participants to categorize themselves on a 4-Likert scale, where 1 is definitely a morning person, 2 is mostly a morning person, 3 is mostly an evening person and 4 is definitely an evening person. For this study, participants categorized as 1 or 2 were labelled morning chronotype and participants categorized as 3 or 4 were labelled evening chronotype.

### Functional data acquisition and preprocessing

Baseline structural T1w and resting state functional MRI (rs-fMRI) data were available and used for further analyses. Both structural and functional scans were acquired with a 3T Siemens Skyra scanner and a standard Siemens 32-channel radiofrequency receiver head coil. Parameters for fMRI included: field of view (FOV) = 88 × 88 × 64, voxel resolution = 2.4mm isotropic, flip angle (FA) = 52°, TR/TE = 0.735s/0.039s, total scan duration = 6 minutes, using a multiband acceleration of factor 8, resulting in 490 volumes per subject. Data were acquired with eyes open looking at a fixation cross. Preprocessing of rs-fMRI images was performed using fMRIPrep version 25.2.5 via Apptainer in BIDS format [39]. Preprocessing steps included brain extraction, motion correction, slice timing correction and co-registration of the blood-oxygenation level dependent (BOLD) image to the T1w structural image. Due to the absence of fieldmaps for UK Biobank data, susceptibility distortion correction was applied using the SyN-based SDC method (option --use syn-sdc). The output was specified to individual T1w native space. All other parameters were left at default. Denoising was done via the Nilearn Python toolbox using the confounds file output from fMRIPrep. Hyperparameter decisions were made in line with Jiang et al. (2021), who were also interested in small subcortical nuclei. A 16-parameter nuisance regressor strategy was used, including six motion parameters, their temporal derivatives, as well as mean white matter and cerebrospinal fluid (CSF) signals along with their temporal derivatives. Bandpass filtering between 0.009 and 0.08 Hz was applied. Non-steady state volumes (as described in the confounds file) and volumes exceeding the framewise displacement threshold of 0.5mm were censored prior to denoising. Subjects with less than 240 seconds of retained data were excluded from further analysis.

### Structural data acquisition and segmentations

Segmentation of the regions of interest (ROI) was performed based on T1w structural data, which was acquired with parameters: FOV=256 × 256 × 208, voxel resolution=1-mm isotropic, integrated parallel acquisition technique in-plane acceleration factor=2, prescan normalization = true, total scan duration = 5 minutes. Preprocessing of T1w data was performed with FreeSurfer’s automated recon-all pipeline using version 7.4.1 [41]. This includes classical preprocessing steps such as motion correction, skull-stripping and registration; see Freesurfer documentation for more details (https://surfer.nmr.mgh.harvard.edu/fswiki/recon-all). SCN and PG were segmented using FreeSurfer’s automated segmentation tools (Fig. 1A). The SCN is estimated to be (1.7 × 1.1 × 1.1) mm^3^ ∼ 2.1 mm^3^ and likely sits between voxels, thus segmention at the current 1mm^3^ resolution proves challenging. As such, instead of SCN segmentation, the SCN-containing region, i.e., the anterior-inferior hypothalamus (AIH) was used. This region contains both the SCN and the supraoptic nucleus. The left- and right-AIH labels (IDs 801 and 806), were extracted using FreeSurfer’s hypothalamic segmentation pipeline [42]. The PG (label ID 900) was segmented using FreeSurfer’s PGlands segmentation tool [43]. All segmentations were obtained in each participant’s native space, resulting in marginal variation in ROI sizes across participants. As FreeSurfer outputs reside in a different space than the T1w acquisition parameters produced by fMRIPrep, segmentation masks were first brought into alignment with the native T1w structural image space using FreeSurfer’s mri_label2vol function. Given that the BOLD image has a resolution of 2.4mm isometric while the segmentations are at 1mm isometric, regridding had to be done to bring the masks and the BOLD image into the same resolution. Since the ROIs are anatomically small, resampling the masks down to BOLD resolution resulted in substantial voxel loss; instead, the BOLD time series was upsampled to 1mm isometric mask space. This resampling was carried out using SPM’s image calculator [44]. The outputs for each subject were concatenated using MRtrix3’s mrcat command [45]. The final 4D image resulting from this command was used as input for the subsequent step. To reduce the high-dimensional voxel-wise timeseries within each ROI to a single representative signal suitable for DCM, singular value decomposition (SVD) was applied. Voxels were masked to exclude zero and NaN values, and the resulting voxel-by-time matrix was mean-centred prior to decomposition. The first eigenvariate – the temporal component explaining the greatest variance across voxels – was retained as the ROI timeseries. A sign correction was applied to ensure the first eigenvariate was aligned with the mean signal across voxels. The resulting eigenvariates were used as input for the DCM analysis.

**Figure 1.**
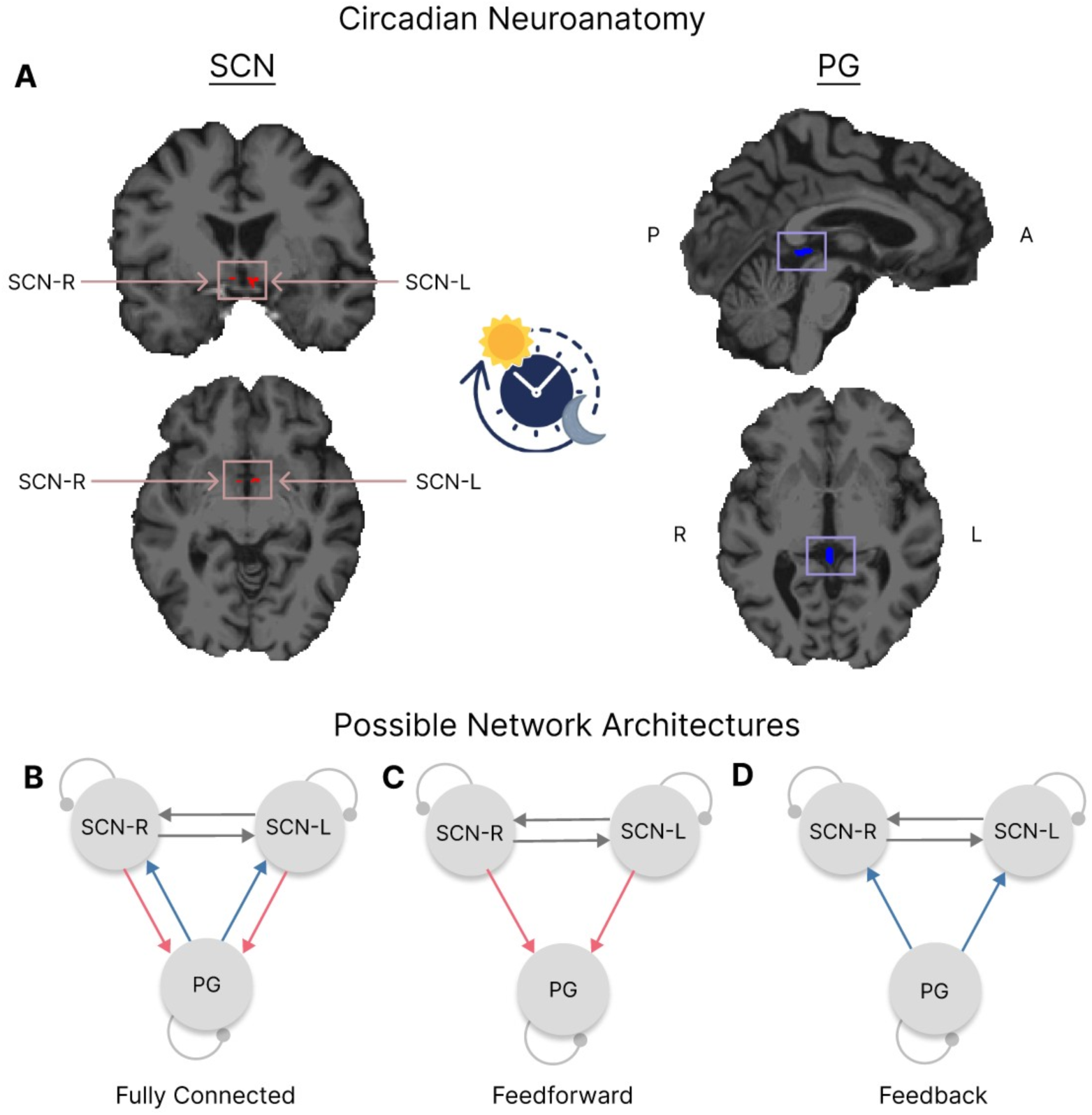
Circadian regions-of-interest and explored DCM model architectures. **A:** The left shows the mask of the SCN-containing anterior-inferior hypothalamic subunit, overlaid in red on the native T1-weighted MRI of an exemplary sample participant. The right shows the pineal gland (PG) mask overlaid in blue over the same brain. A clock icon showing sun and moon separates them to emphasize circadian relevance. **B:** Visualization of the fully connected model for DCM. **C:** Feedforward-only model. **D:** Feedback-only model.

### DCM Specification and Estimation

Dynamic Causal Modelling (DCM) for resting-state fMRI was applied to characterize effective connectivity among the three regions of interest: the left- and right-SCN-containing regions and the PG. Specifically, cross-spectral DCM (CSD) was employed, which was a variant of DCM developed for resting-state fMRI data as it fits a generative model directly to the frequency-domain representation of the observed BOLD signals, allowing estimation of effective connectivity from the spontaneous BOLD fluctuations [46]. For each subject, the DCM was specified using the first eigenvariate time series extracted from each ROI as described in the SVD. TR was set to 0.735 s and TE to 0.039 s. Slice timing delays were set to TR/2 = 0.3675 s for all regions. No external driving inputs (C-matrix) or modulatory connections (B-matrix) were specified, consistent with a resting-state paradigm.

### Candidate Model Space

Four candidate DCM architectures were specified for each subject, differing in their endogenous connectivity (A-matrix): (1) a full model, in which all directed connections between the three regions were modelled; (2) a feedforward-only model, retaining only the ascending SCN-to-PG connections and removing the descending PG-to-SCN connections; (3) a feedback-only model, retaining only the descending PG-to-SCN connections and removing the ascending SCN-to-PG connections (Fig. 1B-D); and (4) a null model, retaining only self-connections (diagonal A-matrix) and removing all inter-regional connectivity. The three lesser-connected models were estimated for all 431 subjects using Bayesian Model Reduction (BMR) from each subject’s fitted full model.

### Bayesian Model Selection

Two random-effects Bayesian Model Selection (BMS) analyses were performed using SPM’s spm_BMS function, chosen because our sample spanned multiple diagnostic groups and random-effects. BMS allows model identity to vary across subjects, making it more appropriate and more robust to outliers as compared to fixed-effects BMS, which assumes a single generative model for the entire group [47]. The full, feedforward-only, and null models were first compared to establish whether inter-regional connectivity was present, and if so, whether it was sufficiently explained by a reduced (feedforward) architecture relative to the fully connected model. Second, the feedforward- and feedback-only models were compared directly, collapsed across diagnoses and chronotypes, to test directionality preference of SCN◊PG connectivity for all subjects collapsed. For this second comparison, a per-subject log Bayes factor difference (dF = F(feedback-only) − F(feedforward-only)) was computed to quantify the relative strength of evidence favoring one direction over the other [48]. For both comparisons, a protected exceedance probability (PXP) threshold of 0.95 was used to declare a winning model. PXP describes the probability that a given model is more frequent than the others, adjusted for the null hypothesis that all models are equally likely.

### Model Fit and Sensitivity

SPM has the spm_dcm_fmri_check function to validate various aspects of model fit, however since ROI extraction was not done through SPM, these checks were run manually in MATLAB following the same formulas used by spm_dcm_fmri_check [49, 50]. The first check, absolute model fit at the region level, was calculated using percent variance explained (R^2^) in the fully connected model. The formula per SPM was 100 × PSS / (PSS + RSS), where PSS is the summed squared magnitude of the model’s predicted cross-spectra (DCM.Hc) and RSS is the summed squared magnitude of the residual cross-spectra (DCM.Rc), summed across all frequencies. This was limited to the fully connected model as this is the only model with independently generated predicted spectra. Reduced models were derived analytically via BMR. A threshold of 25% variance explained per region was used to flag subjects with poor absolute fit for each region, respectively [49]. BMS and subsequent statistical analysis were re-run excluding flagged subjects to confirm robustness.

The second check, the largest absolute extrinsic connection strength, was used to assess basic convergence. Per SPM, if no between-region connection exceeds 0.125 Hz, this would suggest that no connection moved meaningfully away from its zero-mean prior for that subject, indicating either non-convergence or overly noisy data.

The third check, the effective number of parameters estimated, was calculated as the Kullback-Leibler divergence between posterior and prior distributions over all parameters, normalized by the log of the number of observations. This reflects how far model estimation moved the posterior away from the prior across the full model (i.e., whether a subject’s fit could simply reflect prior assumptions instead of evidence within the data). Per SPM, this is flagged if the effective number of parameters estimated is less than or equal to 1.

To address the possibility that model comparisons reflect solely prior assumptions rather than evidence in the data, a prior-variance sensitivity analysis was completed using MATLAB [50]. The prior variance of the retained (non-zeroed) SCN-Pineal connections in each reduced model (denoted pC in the DCM output) was scaled by three multipliers for comparison: 0.5x, 1x (default), and 2x. For each multiplier, the reduced model‘s log evidence was recalculated using spm_log_evidence, reusing the same posterior estimates from the fully-connected model fit (no re-estimation). Following this, group-level BMS was computed for each multiplier using the same Morning/Evening chronotype population defined elsewhere. If all three multipliers resulted in the same model being favoured in this population, the finding was considered to be robust.

### Statistics

All statistical analyses were performed in MATLAB [50]. An alpha level of *p* < .05 was used to determine significance throughout and outliers were not excluded from any analyses. Demographic characteristics were compared across the four diagnostic groups (BD, MDD, PY, HC) using one-way ANOVAs for continuous variables and Pearson’s χ² tests of independence for categorical variables. For χ² tests that reached significance, standardized (adjusted) residuals were computed post-hoc for each cell to identify group(s) driving the effect with a threshold of |*z*| > 1.96, corresponding roughly *p* < .05. The independent effects of diagnosis and chronotype on dF were tested with ANCOVAs fit using MATLAB’s fitlm function. Covariates in both ANCOVAs included age, sex, handedness, acquisition time, and when applicable, medication class (antidepressant, lithium, anticonvulsant mood stabilizer, antipsychotic, and other psychiatric medication). See Tahedl et al. (2026) for further details on medication classification.

The interaction effect of diagnosis and chronotype on dF was tested using a separate ANCOVA. Significant interactions were investigated using post-hoc parametric linear contrast testing of chronotype simple effects within each diagnostic group. Contrast testing was carried out using the coefficient estimates and covariance matrix of the fitted interaction model, implemented with MATLAB’s fitlm. For the omnibus ANCOVA tests, an uncorrected alpha level of p < .05 was used. Post-hoc contrast results were familywise error rate (FWER) corrected using the Bonferroni method for comparing across four estimates. Covariates were the same as described above.

## Results

### Participants

Participants were subdivided into four diagnostic groups: HC = 136 (81 morning, 55 evening chronotypes), BD = 78 (50 morning, 28 evening chronotypes), MDD = 154 (74 morning, 80 evening chronotypes) and PY = 63 (30 morning, 33 evening chronotypes). Table 1 shows the complete participant demographics. One-way ANOVAs across groups suggested no differences in mean age (*F*_3,427_ = 0.998, *p* = 0.393), time of day of scan (*F*_3,427_ = 0.795, *p* = .497), or education score (*F*_3,387_ = 0.291, *p* = .832). χ^2^ demonstrated a significant difference in frequency distributions for chronotype (χ^2^_3_ = 8.09, *p* = .044), where post hoc tests showed more evening chronotypes in the MDD group than expected compared to the other three groups (*z*[MDD×EV] = 2.01, *p* < .05). There were no significant differences from χ^2^ for handedness. Lastly, χ^2^ had significant differences across groups for sex (χ^2^_3_ = 14.92, *p* .002), where post hoc showed that there were significantly more females in the MDD group compared to others (*z*[MDD×F] = 3.85, *p* < .01).

**Table 1.** Demographics of the Study Population.

| Variable | Study Group |  |  |  | Statistics |  |  |
| --- | --- | --- | --- | --- | --- | --- | --- |
| | HC,<br>n = 136 | BD,<br>n = 78 | MDD,<br>n = 154 | PY,<br>n = 63 | ANOVA<br>*F* | $\chi^2$ | *p*-value<br>/Post Hoc<br>SDR |
| Age, Years<br>[mean ± SD] | 53.4 ±<br>7.33 | 54.6 ±<br>7.27 | 53.9 ±<br>7.62 | 52.6 ±<br>7.33 | $F_{3,427} =$<br>0.998 | | $p = .393$ |
| Chronotype<br>[Morning/Evening] | 81/55 | 50/28 | 74/80 | 30/33 | | $\chi^2_3 =$<br>8.09 | $p = .044$<br>$z(\text{MDD} \times \text{EV}) = 2.01$<br>* |
| Time of Day of<br>Acquisition (24-hour<br>clock, decimal hours)<br>[mean ± SD] | 13.5 ±<br>3.29 | 13.9 ±<br>3.21 | 13.3 ±<br>2.95 | 13.5 ±<br>3.07 | $F_{3,427} =$<br>0.795 | | $p = .497$ |
| Education Score<br>[mean ± SD] | 12.6 ±<br>12.80 | 13.1 ±<br>17.18 | 13.9 ±<br>14.57 | 14.6 ±<br>13.50 | $F_{3,387} =$<br>0.291 | | $p = .832$ |
| Handedness<br>[Right/Left/<br>Ambidextrous] | 129/6/1 | 67/8/3 | 137/16/1 | 59/3/1 | | $\chi^2_6 =$<br>9.74 | $p = .136$ |
| Sex<br>[Female/Male] | 76/60 | 42/36 | 114/40 | 35/28 | | $\chi^2_3 =$<br>14.92 | $p = .002$<br>$z(\text{MDD} \times \text{F}) = 3.85$ * |
| Antidepressant Intake<sup>a</sup><br>[Yes/No] | 1/135 | 23/55 | 154/0 | 17/46 | NA | NA |  |
| Lithium Intake<sup>b</sup><br>[Yes/No] | 0/136 | 15/63 | 1/153 | 1/62 | NA | NA |  |
| Anticonvulsant Mood<br>Stabilizer Intake<sup>c</sup><br>[Yes/No] | 0/136 | 16/62 | 0/154 | 1/62 | NA | NA |  |
| Antipsychotic Intake<sup>d</sup><br>[Yes/No] | 0/136 | 9/69 | 0/154 | 10/53 | NA | NA |  |
| Other Psychiatric<br>Medication Intake<sup>e</sup><br>[Yes/No] | 0/136 | 3/75 | 1/153 | 1/62 | NA | NA |  |

ANOVAs were performed to test differences in age, years of education, and time of day of acquisition between the 4 diagnosis groups; Pearson's $\chi^2$ tests were performed to test differences in chronotype, handedness, and sex frequencies between the 4 diagnosis groups. For significant $\chi^2$ tests, we investigated standardized residuals post hoc to determine the differing group(s), assuming a significance threshold of $|z| > 1.96 \approx p < .05$ . $\chi^2$ tests were not performed to compare medication frequencies between groups, as medication status is closely tied to diagnosis by clinical definition and several expected cell counts would be too small for valid inference.

### Bayesian Model Selection Results

The first BMS, comparing the fully connected, feedforward-only and null models, resulted in overwhelming favor of the fully connected model. The full model attained a posterior probability of 97.8%, with a PXP of 1.00, while the corresponding values for the reduced and null models were effectively zero for both metrics. The second BMS, comparing the feedforward- and feedback-only models resulted in favor of the feedforward-only model, with posterior probability 70% for feedforward-only and 30% for feedback-only (PXP = 1.00) (Fig. 2).

**Figure 2.**
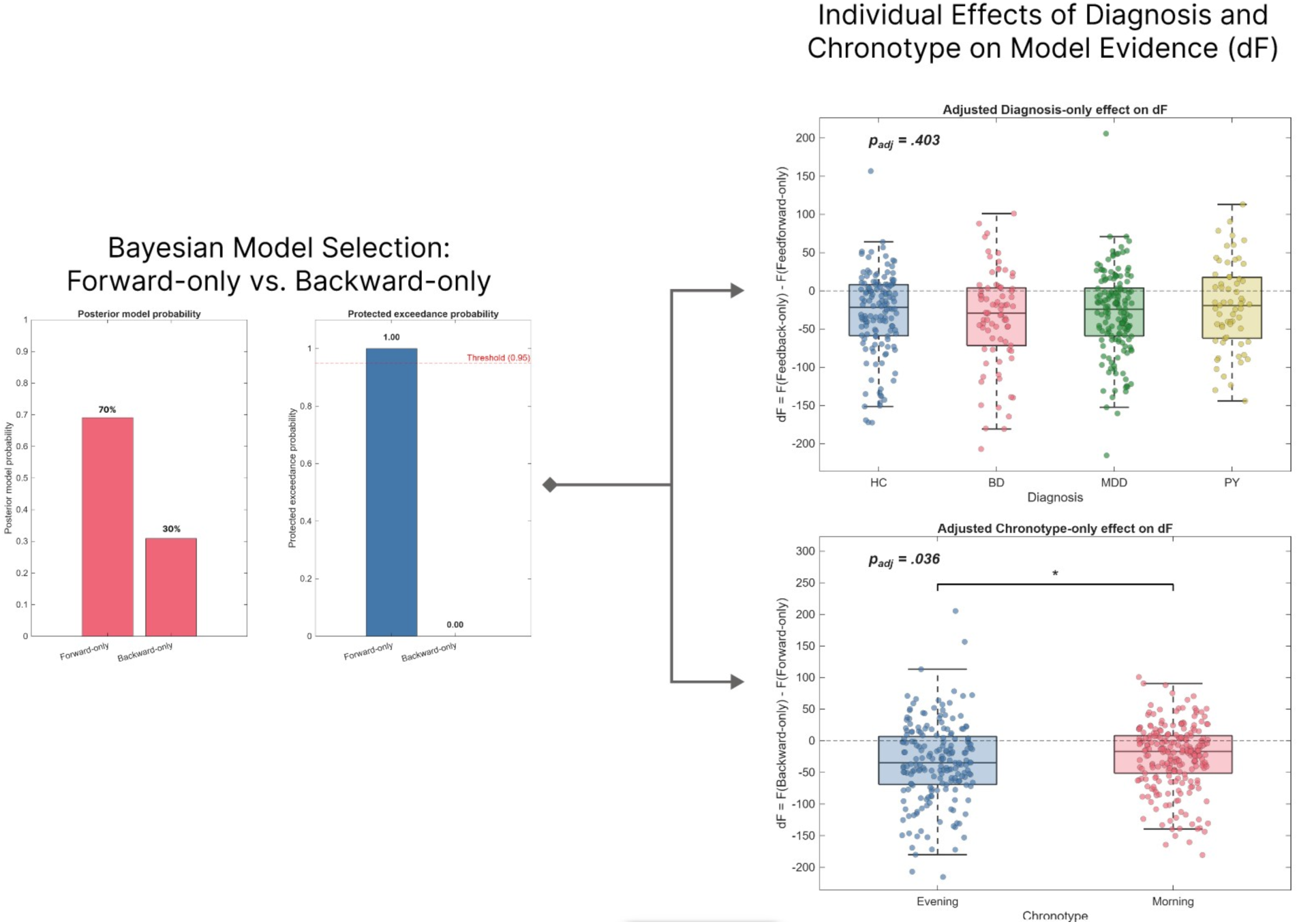
Bayesian Model Selection comparing feedforward- and feedback-only architectures and exploratory analysis comparing preference to diagnosis and chronotype independently. * *p* < 0.05.

### Model Fit and Sensitivity Results

After running the percent variance (R^2^) per region test with a threshold of 25%, 3/431 subjects did not meet the threshold; all in the PG (Supplementary Tab. 1). The feedforward versus feedback BMS and interaction analysis of diagnosis-by-chronotype were repeated excluding the 3 poor fitting subjects – results remained unchanged (Supplementary Tab. 1), thus subjects were not excluded from primary analysis.

For both the second and third check, the largest absolute extrinsic connection strength (threshold to flag < 0.125 Hz) and the effective number of parameters estimated (threshold to flag ≤ 1), respectively, no subjects were flagged (Supplementary Tab. 1).

For the final prior-variance sensitivity check, we found with all three multipliers (0.5, 1, 2), the same model (feedforward) was favoured, indicating the finding is robust to the specific prior variance assumed (Supplementary Tab. 1).

### The effect of diagnosis and chronotype on feedback preference

Subsequent ANCOVA examining the independent effect of diagnosis on feedback preference (dF) showed no significance (*p* = .403, Fig. 2). Conversely, the independent effect of chronotype showed significance (*p* = .036, Fig. 2). The interaction effect of diagnosis and chronotype was statistically significant (*p* = <.001) and post-hoc *t*-tests revealed significance only within the BD group (*t*_413_ = −4.40, *p* = <.001) after correcting for multiple comparisons (Fig. 3 and Supplementary Tab. 1). PY showed a trend with uncorrected *p* = 0.05 (t_413_ = 1.96). Overall, BD evening presented a significantly more negative dF than BD morning, whereas PY presented the reverse, with PY morning presenting the more negative dF, though PY differences did not reach corrected statistical significance.

**Figure 3.**
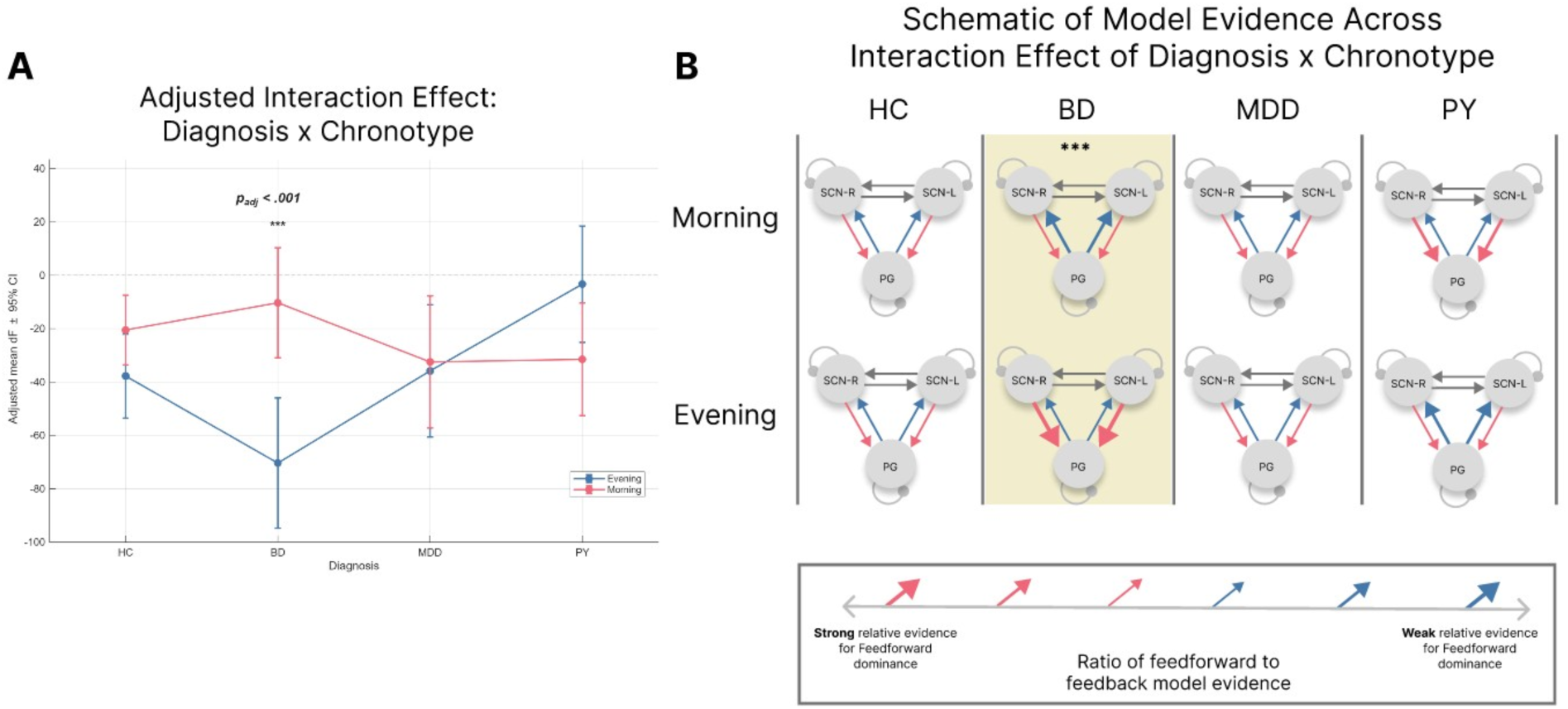
Interaction analysis of the diagnosis-by-chronotype effect on dF. **A:** Interaction plot depicting the effect. *** p < .001. **B:** Visualization depicting ratio of model evidence strength for all possible diagnosis x chronotype combinations. Arrow thickness and color correspond to the strength of relative model evidence for the feedforward model.

## Discussion

In the present study, we tested three a priori hypotheses concerning directed effective connectivity between the SCN-containing anterior-inferior hypothalamic subunit and the PG using DCM of resting-state fMRI data in a large transdiagnostic psychiatric cohort. Bayesian model selection showed a preference for a fully-connected bidirectional model. Despite this preference, model evidence favored feedforward (SCN◊PG) over feedback (PG◊SCN) connectivity. Evening chronotype was associated with greater feedforward dominance. Qualified by a significant diagnosis-by-chronotype interaction, that preference was strongest in the BD evening chronotypes, with no comparable evening-morning difference in other groups.

Bayesian model selection preferred a fully-connected architecture over reduced and null models, meaning a model with both SCN◊PG and PG◊SCN connections accounted for the resting-state cross-spectra better than feedforward-only or disconnected alternatives. This is an inference from BOLD effective connectivity and should not be read as direct evidence of the anatomical loop. Classically, control of pineal melatonin synthesis has been viewed as predominantly feedforward: the SCN exerts tonic daytime inhibition of sympathetic outflow and releases this inhibition at night, permitting norepinephrine-driven melatonin production [19–23]. The subsequent identification of high-affinity MT1 and MT2 receptors on SCN neurons, and human melatonin phase-response curves, already support a functional feedback limb through which melatonin can inhibit SCN firing and phase-shift the master clock [24, 25]. Preference for the fully connected model is consistent with that architecture being estimable from rs-fMRI, while the feedforward direction still dominates when ehte two one-way models are compared directly.

The overall dominance of feedforward models is consistent with the SCN’s established role as the primary circadian pacemaker [51, 52]. Higher feedforward dominance (greater relative evidence for SCN◊PG vs. PG◊SCN) can be interpreted as relatively stronger or less flexible SCN gating of the pineal gland [35, 53], with correspondingly reduced capacity for melatonin-mediated reciprocal fine-tuning of circadian phase or amplitude [54]. Evening chronotype was associated with greater feedforward dominance across the sample. That association was not uniform across diagnoses: it was qualified by the interaction reported above. This is compatible with previous evidence that evening preference confers risk for mood instability, delayed phase, and poorer clinical outcomes across psychiatric disorders [8, 9].

The diagnosis-by-chronotype interaction was driven specifically by BD evening chronotypes, who showed the most extreme feedforward dominance. No analogous pattern emerged in MDD, PY, or HC. This result converges with our prior structural finding of larger volume in the same SCN-containing anterior-inferior hypothalamic subunit selectively in BD evening versus morning chronotypes [13]. Convergence across modalities strengthens the interpretation that the combination of BD diagnosis and evening chronotype constitutes a biologically distinct high-risk subtype [55, 56]. On that reading, BD can be framed as a disorder in which circadian regulation is particularly vulnerable: when it coincides with an evening chronotype, relative model evidence shifts further toward SCN◊PG dominace. That is consistent with emerging consensus that circadian disruption is a core feature of BD pathophysiology [8, 57, 58].

The results support including a simple chronotype assessment (e.g., a single-item preference question or the Morningness–Eveningness Questionnaire) in BD patient care [59]. Whether a shift in relative evidence from feedforward toward feedback coupling tracks response to lithium or chronotherapy is not known and would require a longitudinal or interventional design [60, 61]. Prior multimodal work reported lithium-associated attenuation of certain glial-mitochondrial and structural signatures [14]. Whether lithium is associated with greater relative evidence for feedback in this circuit remains open. These observations align with emerging consensus to integrate circadian measures more systematically into BD treatment guidelines [59, 61].

Several constraints follow from the method and the sample. Cell sizes within the BD morning and evening subgroups were modest, which limits power for finer contrasts. Cross-spectral DCM estimates effective connectivity from BOLD and rests on a closed set of generative models [34, 46, 62]. Bayesian model selection identifies the best model among those specified. It does not however validate that model against the “true” circuit, and a winning architecture can still fit one node poorly [63]. That concern is relevant here: the pineal lies outside the blood-brain barrier, is often calcified in this age range, and occupies few native voxels at the present resolution. We therefore report region-wise variance explained for the fully connected model rather than a single whole-model fit. Three of 431 participants fell below a 25% R^2^ threshold, all in the PG. Excluding them did not change the BMS or the diagnosis-by-chronotype interaction, so they were retained in the primary analysis. R^2^ indexes agreement in spectral power, not spectral shape, so a high value cannot fully exclude a qualitatively poor fit. No participant failed the two SPM convergence checks (largest absolute extrinsic connection < 0.125 Hz, effective number of parameters <= 1), indicating that between-region estimates left the prior and that posteriors were not simply the priors rewritten. A prior-variance sensitivity analysis of the retained SCN-PG connections (0.5x, 1x, 2x default) left the feedforward-versus-feedback ordering unchanged. Residual physiological noise cannot be fully excluded [64, 65]. The SCN-containing anterior-inferior hypothalamic subunit also includes neighboring nuclei (e.g., the supraoptic nucleus and parts of the preoptic area), so osmotic, thermoregulatory, or orexinergic signals may contribute [42]. Moreover, the design is cross-sectional and cannot establish whether connectivity differences precede illness onset or vary with mood state [66].

Despite these constraints, the study benefits from a large transdiagnostic sample, Bayesian model comparison, control for key confounds, and multimodal consistency with prior structural findings. Collectively, the results position BD evening chronotype as a biologically-distinct entity at the level of circadian effective connectivity. If replicated, they support the incorporation of low-burden chronotype assessment into routine and personalized BD patient care. Whether directed SCN-PG effective connectivity can serve as a marker of treatment response is a question for longitudinal and interventional work, not a claim of the present data. Next steps include higher-resolution imaging combined with simultaneous melatonin sampling, interventional designs testing whether modulation of the feedback limb improves clinical outcomes, and longitudinal studies in at-risk individuals to determine whether exaggerated feedforward dominance precedes full illness onset.

## CRediT authorship contribution statement

**IM:** Conceptualization, Data curation, Formal analysis, Methodology, Visualization, Writing – original draft, Writing – review & editing. **JR:** Formal analysis, Validation, Writing – review & editing. **SK:** Validation, Writing – review & editing. **ES**: Funding acquisition, Writing – review & editing. **CC:** Conceptualization, Methodology, Writing – review & editing. **GC:** Conceptualization, Methodology, Writing – review & editing **PH:** Conceptualization, Funding acquisition, Supervision, Writing – review & editing**. MT:** Conceptualization, Data curation, Methodology, Supervision, Writing – original draft, Writing – review & editing.

## Acknowledgments

MT is supported by a project grant from the Hans und Marianne Schwyn-Stiftung and by a Filling-the-Gap fellowship by the Faculty of Medicine, University of Zurich. We wish to thank Imre Kertesz and the rest of the Translational Neuromodeling Unit at ETH Zurich for inspiring this work.

## Data availability

The data used in this study were obtained from the UK Biobank under approved application access. UK Biobank is a large-scale biomedical database that provides controlled access to its data for qualified researchers, subject to application and approval. The data are not publicly available but can be accessed through the UK Biobank repository by researchers who meet the access criteria. The analysis scripts used in this study are available from the corresponding author upon reasonable request.

---

**Supplementary Figure 1.**
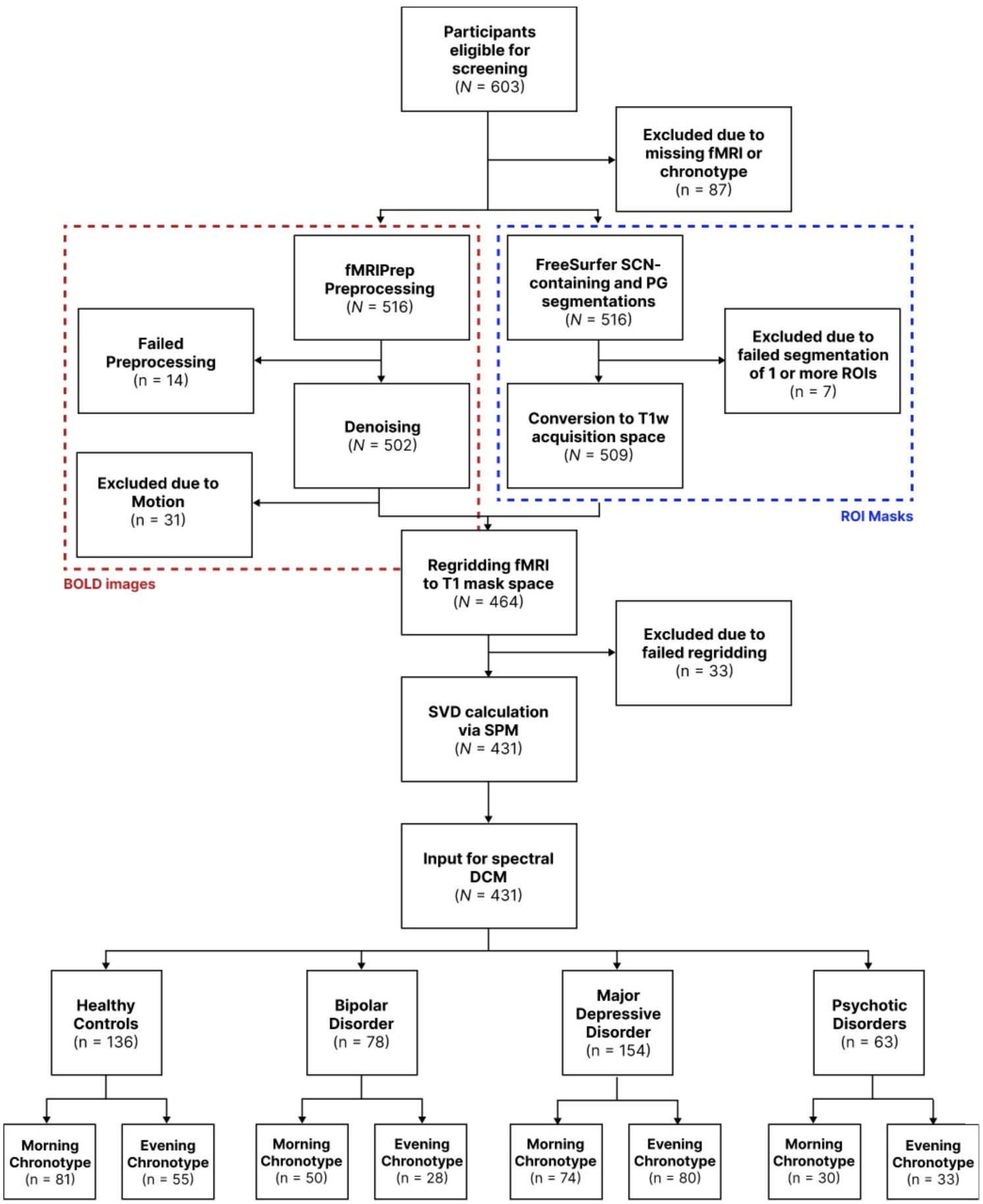
CONSORT diagram depicting flow of participant inclusion/exclusion.

**Supplementary Table 1.** Assessment of Absolute Model Fit and Prior-Variance Sensitivity.

| Percent variance explained ($R^2$) from fully connected model (N = 431) | | | | | | |
| --- | --- | --- | --- | --- | --- | --- |
| | Mean | SD | Median | Min | Max | n flagged below 25% $R^2$ (%) |
| Whole model, % | 84.4 | 9.1 | 87.6 | 32.3 | 94.3 | 0 (0.0%) |
| SCN-L, % | 89.9 | 5.9 | 91.4 | 35.5 | 96.7 | 0 (0.0%) |
| SCN-R, % | 89.3 | 6.1 | 91.0 | 39.6 | 97.0 | 0 (0.0%) |
| PG, % | 85.3 | 13.0 | 90.0 | 19.9 | 96.2 | 3 (0.7%) |
| BMS full sample versus excluding 3 flagged PG poor fit |  |  |  |  |  |  |
|  | Full sample (N = 431) |  |  | Excluding poor fit (N=428) |  |  |
| Feedforward-Feedback BMS, posterior probability (FF) | 0.7013 |  |  | 0.6991 |  |  |
| Feedforward-Feedback BMS, posterior probability (FB) | 0.2987 |  |  | 0.3009 |  |  |
| Feedforward-Feedback BMS, PXP (FF) | 1.00 |  |  | 1.00 |  |  |
| Interaction ANCOVA: Diagnosis × Chronotype, p | 0.0001 | | | 0.0001 | | |
| BD evening-vs-morning simple effect, estimate | -60.0821 |  |  | -60.0480 |  |  |
| BD evening-vs-morning simple effect, $p_{FWER}$ | 0.0001 | | | 0.0001 | | |
| Additional per-subject diagnostics |  |  |  |  |  |  |
|  | Mean | SD | Median | Min | Max | n flagged |
| Largest extrinsic connection strength, Hz (flag < 0.125) | 0.752 | 0.297 | 0.773 | 0.127 | 1.578 | 0 |
| Effective number of parameters estimated (flag $\leq 1$) | 86.6 | 22.46 | 88.56 | 34.36 | 164.58 | 0 |
| Prior-variance sensitivity: Feedforward-feedback BMS across prior variance multipliers |  |  |  |  |  |  |
| Multiplier | Posterior Probability (FF) |  | Posterior Probability (FB) |  | PXP (FF) | % subjects favouring FB |
| 0.5 | 0.6512 |  | 0.3488 |  | 1.00 | 34.88% |
| 1 (default) | 0.7013 |  | 0.2987 |  | 1.00 | 29.87% |
| 2 | 0.7110 |  | 0.2890 |  | 1.00 | 28.90% |

SCN-L = Suprachiasmatic nucleus left. SCN-R = Suprachiasmatic nucleus right. PG = Pineal gland. BMS = Bayesian Model Selection. FF = Feedforward. FB = Feedback. PXP = protected exceedance probability. Estimate = covariate-adjusted mean difference in dF (Evening – Morning). dF = F(Backward-only) – F(Forward-only). $p_{FWER}$ = Bonferroni-corrected p-value ( $\times 4$ diagnosis groups).

**Supplementary Table 2.** Statistical Assessment of an Interaction Effect of Diagnosis and Chronotype on Relative Model Evidence for Feedforward-versus Feedback-Only SCN–Pineal Connectivity.

| Interaction Model |  |  |  |  |
| --- | --- | --- | --- | --- |
|  | Sum Square | Mean Square | F<sub>Dof</sub> | p-value |
| Diagnosis | $1.2 \times 10^4$ | $3.8 \times 10^3$ | $F_{3,413} = 1.26$ | .289 |
| Age | $1.0 \times 10^4$ | $1.0 \times 10^4$ | $F_{1,413} = 3.31$ | .070 |
| Chronotype | $1.6 \times 10^4$ | $1.6 \times 10^4$ | $F_{1,413} = 5.33$ | .021* |
| Sex | $5.5 \times 10^3$ | $5.5 \times 10^3$ | $F_{1,413} = 1.81$ | .180 |
| Handedness | $1.1 \times 10^4$ | $5.3 \times 10^3$ | $F_{2,413} = 1.74$ | .177 |
| Acquisition Time | $6.6 \times 10^1$ | $6.6 \times 10^1$ | $F_{1,413} = 0.02$ | .883 |
| Antidepressant Intake | $9.9 \times 10^2$ | $9.9 \times 10^2$ | $F_{1,413} = 0.32$ | .570 |
| Lithium Intake | $1.6 \times 10^1$ | $1.6 \times 10^1$ | $F_{1,413} = 0.01$ | .943 |
| Anticonvulsant Mood Stabilizer Intake | $1.9 \times 10^0$ | $1.9 \times 10^0$ | $F_{1,413} = 0.00$ | .980 |
| Antipsychotic Intake | $3.8 \times 10^3$ | $3.8 \times 10^3$ | $F_{1,413} = 1.24$ | .267 |
| Other Psychiatric Medication Intake | $6.6 \times 10^3$ | $6.6 \times 10^3$ | $F_{1,413} = 2.14$ | .144 |
| Diagnosis x Chronotype | $6.6 \times 10^4$ | $2.2 \times 10^4$ | $F_{3,413} = 7.13$ | < .001*** |
| Residuals | $1.3 \times 10^6$ | $3.1 \times 10^3$ | NA (*df* = 413) | NA |
| Post-Hoc: Evening vs. Morning within Diagnosis |  |  |  |  |
|  | Estimate | SE | t ratio<sub>Dof</sub> | p<sub>FWER</sub> |
| Bipolar Disorder | -60.08 | 13.64 | $t_{413} = -4.40$ | < .001*** |
| Healthy Control | -17.25 | 9.78 | $t_{413} = -1.76$ | .314 |
| Major Depressive Disorder | -3.36 | 9.16 | $t_{413} = 0.37$ | 1.000 |
| Psychotic Disorders | 28.19 | 14.35 | $t_{413} = 1.96$ | .201 |

